# Procalcitonin for Antimicrobial Stewardship Among Cancer Patients Admitted with COVID-19

**DOI:** 10.1101/2022.07.13.22277580

**Authors:** Hiba Dagher, Anne-Marie Chaftari, Patricia Mulanovich, Ying Jiang, Ray Hachem, Alexandre E. Malek, Jovan Borjan, George M. Viola, Issam Raad

## Abstract

**Background:** Procalcitonin (PCT) has been used to guide antibiotic therapy in bacterial infections. We aimed to determine the role of PCT in decreasing the duration of empiric antibiotic therapy among cancer patients admitted with COVID-19.

**Methods:** This retrospective study included cancer patients admitted to our institution for COVID-19 between March 1, 2020, and June 28, 2021, with a PCT test done within 72 hours after admission. Patients were divided into 2 groups: PCT <0.25 ng/ml and PCT ≥0.25 ng/ml. We assessed pertinent cultures, antibacterial use, and duration of empiric antibacterial therapy.

**Results:** The study included 530 patients (median age, 62 years [range, 13-91]). All the patients had ≥1 culture test within 7 days following admission. Patients with PCT <0.25 ng/ml were less likely to have a positive culture than were those with PCT ≥0.25 ng/ml (6% [20/358] vs 17% [30/172]; p<0.0001). PCT <0.25 ng/ml had a high negative predictive value for bacteremia and 30-day mortality. Patients with PCT <0.25 ng/ml were less likely to receive intravenous (IV) antibiotics for >72 hours than were patients with PCT ≥0.25 ng/ml (45% [162/358] vs 69% [119/172]; p<0.0001). Among patients with PCT <0.25 ng/ml and negative cultures, 30-day mortality was similar between those who received IV antibiotics for ≥72 hours and those who received IV antibiotics for shorter durations (2% [2/111] vs 3% [5/176], p=0.71).

**Conclusions:** Among cancer patients with COVID-19, PCT level <0.25 ng/ml is associated with lower likelihood of bacterial co-infection and greater likelihood of a shorter antibiotic course. In patients with PCT level <0.25 ng/ml and negative cultures, an antibiotic course of > 72 hours is unnecessary. PCT could be useful in enhancing antimicrobial stewardship in cancer patients with COVID-19.

## Introduction

Many factors predicting the outcome and prognosis of coronavirus disease 2019 (COVID-19) have been identified. These factors have proved valuable for determining prognosis and have guided the treatment of patients at risk for severe COVID-19. Procalcitonin (PCT) is a biomarker that has served as an indicator for bloodstream infections and has been used as a guide to antimicrobial management in sepsis and bacterial infections in the general population^1-4^ and in cancer patients with and without neutropenia^5-8^. Randomized trials have shown PCT to be useful in guiding decisions regarding antimicrobial therapy for lower respiratory tract infections^9-11^. Several PCT cut-off values have been evaluated and used in different treatment algorithms. PCT cut-off values of 0.25 ng/ml and 0.5 ng/ml have been adopted for critically ill patients in the intensive care unit (ICU)^12^, neutropenic patients^2,3,8^, and patients with lower respiratory tract infections^13^.

In patients with coronavirus disease 2019 (COVID-19), elevated PCT levels and elevated levels of other inflammatory markers have been associated with more severe COVID-19 both in the general population^5,14-17^ and in cancer patients^1,18^.

Bacterial co-infections may not be prevalent in patients with COVID-19^19^. However, because of the similarity in signs and symptoms between bacterial co-infections and COVID-19 and the difficulty of ruling out a bacterial infection in patients presenting with COVID-19 pneumonia, empirical treatment with antimicrobials is often initiated in patients with COVID-19 without a confirmed bacterial co-infection^19^. This practice may lead to an emergence of antimicrobial resistance, undesirable adverse events, and increase costs ^2,3^. One study showed that the use of antibiotics in patients with COVID-19 with a PCT level >0.25 ng/ml and with a low suspicion of bacterial infection did not improve clinical outcome^20^. Little to no data have been published regarding PCT for antimicrobial stewardship among cancer patients with COVID-19.

Given the widespread use of empiric antibiotics in cancer patients admitted for COVID-19, we evaluated the role of PCT in decreasing the duration of empiric antimicrobial therapy in this patient population.

## Methods

We conducted a retrospective study of cancer patients who were admitted to The University of Texas MD Anderson Cancer Center between March 1, 2020, and June 28, 2021, for COVID-19 and had a serum PCT level measured within 72 hours after admission. Patients were divided into 2 groups: PCT level < 0.25 ng/ml and PCT level ≥ 0.25 ng/ml. This cut-off is conventionally suggested and has been used in different algorithms ^11-13^.

We reviewed the patients’ electronic medical records and collected data pertinent to demographics (age, sex, and race and ethnicity), type of cancer (hematological malignancy vs solid tumor), cancer status (active vs in remission), active cancer therapy, co-morbidities, tobacco use, and presence of pneumonia. We assessed laboratory test results, including absolute neutrophil count, PCT level, documented bacterial infections, and sources of cultures. We also extracted data on oxygen saturation, requirement for oxygen supplementation, need for and duration of intravenous (IV) antibiotic therapy, ICU admission, and 30-day mortality after COVID-19 diagnosis.

Our study was approved by the Institutional Review Board of MD Anderson Cancer Center, and a waiver of informed consent was obtained.

We compared the clinical characteristics and outcomes of the patients in the PCT < 0.25 ng/ml and PCT ≥ 0.25 ng/ml groups. We used the χ2 or Fisher’s exact test, as appropriate, to compare categorical variables. We used Wilcoxon rank-sum tests to compare continuous variables because of the deviation of the data from the normal distribution. We assessed negative predictive values of PCT levels for the prediction of the various outcomes. We also estimated the relative risks of various outcomes for a patient with PCT ≥ 0.25 ng/ml. All tests were 2-sided at a significance level of .05. The statistical analyses were performed using SAS version 9.3 (SAS Institute Inc, Cary, NC).

## Results

We identified 530 patients, of whom 358 (68%) had a PCT level < 0.25 ng/ml and 172 (32%) had a PCT level ≥ 0.25 ng/ml. Patients in the 2 PCT groups were similar in terms of age, sex, race and ethnicity, type and status of cancer, and active cancer therapy (Table 1). The rate of an absolute neutrophil count < 1000/µl was 9% in both groups; however, the rate of an absolute lymphocyte count < 1000/µl was lower in patients with PCT < 0.25 ng/ml (63% vs 75%; p=0.009).

**Table 1.** Comparison of hospitalized cancer patients with COVID-19 with different PCT levels.^a^

| Characteristic | PCT < 0.25 ng/ml<br>(n=358) | PCT ≥ 0.25 ng/ml<br>(n=172) | *p*-value |
| --- | --- | --- | --- |
| Age, median (range), years | 61 (13-91) | 64 (14-86) | 0.11 |
| Sex, male | 178 (50) | 95 (55) | 0.23 |
| Race and ethnicity |  |  | 0.36 |
| Asian | 14/355 (4) | 5/171 (3) |  |
| Black | 42/355 (12) | 30/171 (18) |  |
| Hispanic | 102/355 (29) | 47/171 (27) |  |
| White | 194/355 (55) | 86/171 (50) |  |
| Other race or ethnicity | 3/355 (1) | 3/171 (2) |  |
| Declined to answer or unknown | 3 | 1 |  |
| Type of cancer |  |  | 0.79 |
| Hematological malignancy only | 142 (40) | 63 (37) |  |
| Solid tumor only | 195 (54) | 99 (58) |  |
| Both of above | 21 (6) | 10 (6) |  |
| Status of cancer |  |  | 0.90 |
| Active | 315 (88) | 152 (88) |  |
| No evidence of disease | 43 (12) | 20 (12) |  |
| Active cancer therapy within 30 days | 118 (33) | 58 (34) | 0.86 |
| Chemotherapy received | 272 (76) | 121 (70) | 0.16 |
| Smoking status |  |  | 0.29 |
| Never smoker | 223/354 (63) | 96/165 (58) |  |
| Current or former smoker | 131/354 (37) | 69/165 (42) |  |
| Unknown | 4 | 7 |  |
| Chronic kidney disease | 114/327 (35) | 89/168 (53) | < 0.001 |
| Asthma | 44/327 (13) | 23/168 (14) | 0.94 |
| Chronic obstructive pulmonary disease | 59/327 (18) | 34/168 (20) | 0.55 |
| Congestive heart failure | 46/327 (14) | 33/168 (20) | 0.11 |
| Diabetes mellitus | 164/327 (50) | 83/168 (49) | 0.87 |
| Coronary artery disease | 12/327 (4) | 3/168 (2) | 0.25 |
| Hypertension | 251/327 (77) | 137/168 (82) | 0.22 |
| Venous thromboembolic event | 42/327 (13) | 19/168 (11) | 0.62 |
| Obesity | 37/327 (11) | 20/168 (12) | 0.85 |
| Obstructive sleep apnea | 55/327 (17) | 18/168 (11) | 0.07 |
| ANC < 1000/μl at admission | 33/352 (9) | 16 (9) | 0.98 |
| ALC < 1000/μl at admission | 219/348 (63) | 126/169 (75) | 0.009 |
| Pneumonia | 270/357 (76) | 141 (82) | 0.10 |
| Oxygen supplementation within 72 hours | 191/356 (54) | 118 (69) | 0.001 |
| Culture test performed | 358 (100) | 172 (100) |  |
| Positive bacterial culture | 20 (6) | 30 (17) | < 0.0001 |
| Site of positive culture |  |  |  |
| Blood | 2/20 (10) | 10/30 (33) | 0.09 |
| Lower respiratory tract | 5/20 (25) | 10/30 (33) | 0.53 |
| Wound | 6/20 (30) | 5/30 (17) | 0.31 |
| Urine | 10/20 (50) | 8/30 (27) | 0.09 |
| Transfusion reaction culture | 0/20 (0) | 1/30 (3) | > 0.99 |
| Cerebrospinal fluid | 0/20 (0) | 1/30 (3) | > 0.99 |
| Positive fungal culture | 3 (1) | 8 (5) | 0.007 |
| Viral co-infection | 2 (1) | 0 (0) | > 0.99 |
| Duration of hospital stay, median (IQR), days | 6 (4-10) | 10 (6-18) | < 0.0001 |
| IV antibiotic treatment | 271 (76) | 154 (90) | < 0.001 |
| Duration of IV antibiotic treatment, median (IQR), days | 4 (2-6) | 6 (3-7) | < 0.0001 |
| Duration of IV antibiotic therapy ≥ 72 hours | 162 (45) | 119 (69) | < 0.0001 |
| Duration of IV antibiotic therapy ≥ 7 days | 54 (15) | 54 (31) | < 0.0001 |
| Number of different IV antibiotic treatment, median (IQR) | 2 (1-3) | 2 (2-3) | < 0.001 |
| ICU admission | 51 (14) | 50 (29) | < 0.0001 |
| Duration of ICU stay, median (IQR), days | 1 (1-4) | 3 (1-3) | 0.13 |
| Mortality within 30 days of COVID-19 diagnosis | 20 (6) | 33 (19) | < 0.0001 |

ALC, absolute lymphocyte count; ANC, absolute neutrophil count; IQR, interquartile range.
<sup>a</sup>Values in table are number of patients (percentage) unless otherwise indicated.

Patients with PCT < 0.25 ng/ml were less likely to require oxygen supplementation within 72 hours of admission (54% vs 69%, p=0.001); were less likely to have a positive bacterial culture (6% vs 17%; p<0.0001) from any source, including blood, lower respiratory tract, and urine; and had a lower rate of pneumonia, although the difference was not significant (76% vs 82%; p=0.10). Patients with PCT < 0.25 ng/ml were less likely to receive IV antibiotic therapy than were patients with PCT ≥ 0.25 ng/ml (76% vs 90%; p<0.001). Furthermore, patients with PCT < 0.25 ng/ml had a shorter median duration of IV antibiotic therapy (4 days vs 6 days; p<0.0001) and were less likely to receive antibiotics for ≥ 72 hours compared to patients with PCT ≥ 0.25 (45% vs 69%; p<0.0001) (Table 1). Similar results were found among patients with negative culture results: those with PCT < 0.25 ng/ml were less likely to receive IV antibiotics for ≥ 72 hours than those with PCT ≥ 0.25 ng/ml (44% vs 67%; p<0.0001). In addition, among patients with PCT < 0.25 ng/ml and negative culture results, those who received a long course of IV antibiotics (≥ 72 hours) and those who received a shorter course had similar 30-day mortality rates (2% vs 3%, p=0.71) (Table 2).

**Table 2.** Treatment and outcomes of hospitalized cancer patients with COVID-19 with PCT < 0.25 ng/ml and negative bacterial cultures.

| Outcome | Duration of IV antibiotic treatment |  | *p*-value |
| --- | --- | --- | --- |
|  | < 72 hours<br>(n=176) | ≥ 72 hours<br>(n=111) |  |
| Duration of hospital stay, median (IQR), days | 5 (3-7) | 7 (5-11) | < 0.0001 |
| Mortality within 30 days of COVID-19 diagnosis, n (%) | 5 (3) | 2 (2) | 0.71 |

Note: Patients with ICU admission during hospitalization and patients who died within 3 days after hospital admission were excluded from analysis.

Compared to patients with PCT ≥ 0.25 ng/ml, patients with PCT < 0.25 ng/ml had shorter median duration of hospital stay (6 days vs 10 days; p<0.0001), lower rate of ICU admission (14% vs 29%; p<0.0001), and lower rate of mortality within 30 days of COVID diagnosis (6% vs 19%; p<0.0001).

PCT < 0.25 ng/ml had a high negative predictive value for bacteremia, 30-day mortality, ICU admission, and IV antibiotic use ≥ 7 days (Table 3).

**Table 3.** Negative predictive value (NPV) of PCT < 0.25 ng/ml and relative risk associated with PCT ≥ 0.25 ng/ml for selected outcomes in hospitalized cancer patients with COVID-19.

| Outcome | NPV of PCT <<br>0.25 ng/ml | 95% Confidence interval | Relative risk of PCT ≥ 0.25<br>ng/ml | 95% Confidence interval |
| --- | --- | --- | --- | --- |
| Positive bacterial culture | 0.94 | 0.92 to 0.97 | 3.12 | 1.83 to 5.34 |
| Use of IV antibiotics | 0.24 | 0.20 to 0.29 | 1.18 | 1.09 to 1.28 |
| Use of IV antibiotics ≥ 72 hours | 0.55 | 0.49 to 0.60 | 1.53 | 1.31 to 1.78 |
| Use of IV antibiotics ≥ 7 days | 0.85 | 0.81 to 0.88 | 2.08 | 1.50 to 2.90 |
| ICU admission | 0.86 | 0.82 to 0.89 | 2.04 | 1.45 to 2.88 |
| Death within 30 days after COVID-19<br>diagnosis | 0.94 | 0.92 to 0.97 | 3.43 | 2.03 to 5.80 |

PCT level ≥ 0.25 ng/ml was associated with elevated relative risk for 30-day mortality, positive bacterial culture, IV antibiotic use ≥ 7 days, and ICU admission (Table 3).

## Discussion

In this study of cancer patients admitted for COVID-19, we found that PCT level < 0.25 ng/ml was associated with a lower rate of bacterial co-infection, shorter hospital stay, shorter duration of IV antibiotics, and lower 30-day mortality. We also found that among the patients with PCT < 0.25 ng/ml and negative bacterial cultures, 30-day mortality was similar for patients treated with IV antibiotics for ≥ 72 hours and those treated with IV antibiotics for shorter periods.

Our finding that the rate of microbiologically documented bacterial co-infections from any source, including blood, lower respiratory tract, and urine, was lower in patients with PCT < 0.25 ng/ml is consistent with well-established findings that pure viral infections are unlikely to increase PCT levels^21^. Furthermore, both in the general population^1,2,4,9-11^ and in immunocompromised patients^5-8^, patients with low PCT levels are unlikely to have bacterial infections. PCT levels increase in patients with many types of bacterial infections, including bacterial infections of the lower respiratory tract ^22^, bacterial meningitis^23^, acute pyelonephritis^24^, spontaneous bacterial peritonitis^25^, and bloodstream bacterial infections^26^. Our findings regarding PCT levels and the risk of bacterial infection is also consistent with published data on patients with COVID-19^20,27^. In a recent study of patients hospitalized with COVID-19, PCT levels were higher in patients with proven bacterial co-infections: PCT level ≥ 0.25 ng/ml was seen in 69% of patients with proven co-infection, compared to 35% of those with low suspicion of bacterial co-infection (p<0.001)^20^. The low rate of bacterial co-infection in our cancer patients with COVID-19 (about 9%) is also consistent with rates reported in the literature^19,28^.

Another recent study showed that PCT could be abnormally elevated in patients with COVID-19 with no evidence of pneumonia and may result in unnecessary antibiotic administration in such patients^29^. In our current study, IV antibiotics were administered to 90% of patients with PCT ≥ 0.25 ng/ml and 76% of patients with PCT < 0.25 ng/ml (p<0.001). These high rates are similar to rates reported earlier in the pandemic, which ranged from 70% to 90%^30,31^. This high rate of use of IV antibiotics in our cancer patient population could be due to the vulnerability of our immunocompromised patients. The initial PCT level may not have influenced the decision of the treating physician to initiate IV antibiotics in our frail and immunocompromised cancer patient population.

In hospitalized patients with COVID-19, PCT level ≥ 0.25 ng/ml was previously found to be a good predictor of oxygen supplementation, ICU admission, mechanical ventilation, and antibiotic use^20^. Similarly, in our study, cancer patients with higher PCT levels were more likely to require oxygen supplementation within 72 hours of admission, had a higher rate of ICU admission, had a higher 30-day mortality rate, had a longer median duration of hospital stay, and were more likely to receive IV antibiotics.

Our data demonstrate that administering IV antibiotics beyond 72 hours in patients with PCT < 0.25 ng/ml and negative bacterial cultures does not improve outcome and is unnecessary. Thus, just as PCT has been used to de-escalate antibiotic use in the general population ^9,10^, it can be used to de-escalate antibiotic use in cancer patients with COVID-19. We found that patients with PCT < 0.25 ng/ml and negative blood cultures were more likely to receive a short course of antibiotics (< 72 hours) than were patients with PCT ≥ 0.25 ng/ml and negative blood cultures. In addition, we found that among patients with PCT < 0.25 ng/ml and negative blood cultures, patients who received IV antibiotics for < 72 hours had similar 30-day mortality and a significantly shorter hospital stay compared to patients who received IV antibiotics for ≥ 72 hours.

Our findings that PCT < 0.25 ng/ml had a negative predictive value for bacteremia, 30-day mortality, ICU admission, and IV antibiotic use > 7 days are consistent with previously published data from patients with COVID-19^20,32^.

The use of PCT levels to guide antimicrobial therapy decisions has been important in antimicrobial stewardship outside of the COVID-19 pandemic. However, our data suggest that in cancer patients with COVID-19, if the PCT level is < 0.25 ng/ml, there is low suspicion for infection, and if bacterial cultures are negative, PCT could be used as an adjunct to clinical judgment to guide de-escalation of antimicrobials after 72 hours. Incorporating PCT into future algorithms for treatment of patients with COVID-19 could be cost-effective and may decrease antibiotic overuse, which is associated with undesirable adverse events (such as *Clostridium difficile* infection, acute kidney injury, potential allergic reactions, and loss of microbiome diversity) and contributes to the emergence of antibiotic resistance^2,33,34^.

Our study has limitations. First, the retrospective nature of this study may have masked confounding variables. Second, bacterial co-infections may have been overlooked given the limited face-to-face interactions with patients admitted with COVID-19 during the pandemic. Third, antimicrobials were administered empirically at the discretion of the team treating the patient. Finally, this is a single-center study, which limits the generalizability of our results.

## Conclusions

Cancer patients with COVID-19 often receive IV antibiotics despite a low rate of bacterial co-infections. Patients with low PCT levels (< 0.25 ng/ml) are unlikely to have a documented bacterial infection, and they are more likely than patients with higher PCT levels to have a shorter hospital stay, shorter course of IV antibiotics, and a better overall outcome.

In cancer patients with COVID-19 and PCT < 0.25 ng/ml, continuing antibiotics beyond 72 hours (or beyond when the PCT result becomes available, if antibiotic therapy has already been administered for ≥72 hours at that time) does not reduce mortality and is unnecessary. Hence, PCT could be used along with clinical judgment to promote antibiotic stewardship in cancer patients with COVID-19 by reducing the duration of antimicrobial therapy beyond the initial empiric use of systemic antibiotics until PCT results become available.

## Data Availability

These are human subjects of multicenter trials and we are unable to share data because of IRB restrictions from different sites in different countries. The study protocol, statistical analysis plan, lists of deidentified individual data, generated tables and figures will be made available upon request by qualified scientific and medical researchers for legitimate research purposes. Requests should be sent to. Data will be available on request for 6 months from the date of publication. Investigators are invited to submit study proposal requests detailing research questions and hypotheses in order to receive access to these data. The software we used for data analysis is SAS version 9.3 (SAS Institute Inc., Cary, NC), and we have provided this information in Statistical analysis section of the manuscript.

## ACKNOWLEDGMENTS

We thank Ms. Salli Saxton, Department of Infectious Diseases, Infection Control and Employee Health, MD Anderson Cancer Center, Houston, for helping with the submission of the manuscript.

We thank Stephanie Deming, Research Medical Library, MD Anderson Cancer Center, for editing the manuscript.

This research was supported by the National Institutes of Health/National Cancer Institute under award number P30CA016672, which supports MD Anderson Cancer Center’s Clinical Trials Office.

